# Impulse dispersion of aerosols during singing and speaking

**DOI:** 10.1101/2020.07.21.20158832

**Authors:** Matthias Echternach, Sophia Gantner, Gregor Peters, Caroline Westphalen, Tobias Benthaus, Bernhard Jakubaß, Liudmila Kuranova, Michael Döllinger, Stefan Kniesburges

## Abstract

Group singing events have been linked to several outbreaks of infection during the CoVID-19 pandemic, leading to singing activities being banned in many areas across the globe. This link between singing and infection rates supports the possibility that aerosols are partly responsible for person-to-person infection. In contrast to droplets, the smaller aerosol particles do not fall to the ground within a short distance after being expelled by e.g. a singer. Aerosol particles hover and spread via convection in the environmental air. According to the super-spreading theory, choir singing and loud talking (theater and presentations) during rehearsals or performances may constitute a high risk of infectious virus transmission to large numbers of people. Thus, it is essential to define the safety distances between singers in super-spreading situations.

The aim of this study is to investigate the impulse dispersion of aerosols during singing and speaking in comparison to breathing and coughing. Ten professional singers (5 males and 5 females) of the Bavarian Radio Chorus performed 9 tasks including singing a phrase of Beethoven’s 9th symphony, to the original German text. The inhaled air volume was marked with small aerosol particles produced via a commercial e-cigarette. The expelled aerosol cloud was recorded with three high definition TV cameras from different perspectives. Afterwards, the dimensions and dynamics of the aerosol cloud was measured by segmenting the video footage at every time point.

While the median expansion was below 1m, the aerosol cloud was expelled up to 1.4m in the singing direction for individual subjects. Consonants produced larger distances of aerosol expulsion than vowels. The dispersion in the lateral and vertical dimension was less pronounced than the forward direction. After completion of each task, the cloud continued to distribute in the air increasing its dimensions. Consequently, we propose increasing the current recommendations of many governmental councils for choirs or singing at religious services from 1.5m to the front and 1m to the side to a distance between choir singers of 2m to the front and 1.5m to the sides.

## Introduction

Person-to-person transmission of CoVID19 is suspected to occur mainly by direct contact or transmission of small droplets with diameters ≥5 μm and aerosols with diameters ≤ 5 μm(*1*). According to the super-emitter theory(*2*), expulsion of a much larger amount of droplets and aerosols owing to exhalation activities such as extensive and loud speaking, singing, sneezing or coughing poses the highest risk for CoVID19 transmission. With regard to droplets, the distance from the mouth reached by the particles depends on many factors such as loudness (*2*) (*3*)or the type of activity such as vocalizing or articulating consonants. For coughing, particles have been detected at distances up to 8m from the mouth(*4*). After being expelled, large droplets follow ballistic trajectories and descend to the ground quickly and over a short distance(*5-7*). It is from these findings concerning droplets that the most recent recommendations for safe distances between singers in choir rehearsals during the CoVID19 pandemic have been based(*8*).

While the transmission of CoVID19 by droplets is scientifically accepted, until very recently the World Health Organization (WHO) only considered aerosols as an infection transmission route if these particles are generated during supportive treatment or medical interventions such as in- /extubation or tracheostomy(*9, 10*). The WHO now states that transmission by aerosols cannot be ruled out due to recent studies showing mounting evidence for the hypothesis that CoVID19 could spread airborne by aerosols that are generated during speech and especially singing(*11-13*). This possibility that aerosols play a major role in the person-to-person transmission of CoVID19 could explain the fatal infectious transmissions during choir singing in Amsterdam, Berlin and Skagit Country, Washington(*14-16*).

Speaking is thought to produce about three times the absolute amount of aerosol compared to breathing(*17*) and the particle emission rate during different spoken texts increases for phrases that include a higher number of vowels(*18*). Singing, as defined by the sustained vocalization of a vowel, is associated with a much higher rate of aerosol emission, showing ten times higher values than breathing and about three times higher values than speaking(*17*). In a recent pre-print study, the authors observed an even higher production of aerosols with a 4 to 100 times greater amount of aerosols produced by singing compared to speaking(*19*). Therefore, in recent scientific and administrative evaluations, singing has become associated with a much higher risk for person-to-person transmission of infectious diseases, such as CoVID19. Consequently, in certain countries across the globe, singing in choirs and at religious services is now restricted or prohibited(*20, 21*).

Whereas virus-laden droplets are only received when that person is in close proximity to the emitter, aerosols remain hovering and convectively spread in the environmental air. Therefore, aerosols can potentially infect people in two ways: via reception of an impulse emitted from an infected person, or inhalation in a closed room with a virus accumulated virus cloud. In this context, the impulse spreading characteristics of aerosols have not yet been investigated for all exhalation activities in detail. Particularly, the characteristics for singing are yet to be measured. Detailed data of characteristics during speech and singing with the special attention on vowels and consonants could contribute to the understanding of the fatal transmissions in the choir rehearsals mentioned above. Moreover, the extent to which singing only vowels differs from singing a text with potentially heightened consonant production has yet to be considered with respect to the dispersion of aerosols.

This study aims to analyse the impulse dispersion dynamics of aerosols in professional choir singers with the question of how much such dispersion differs between singing a text, singing a vowel or speaking. Furthermore, it has been investigated how much different loudness contributes to the distances reached by the aerosols from the mouth. The measured distances should result in recommendations for safety distances for singing in order to reduce the transmission of infectious diseases, such as CoVID19.

## Material and Methods

After approval from the local ethics committee (no. 20-395), 10 professional full-time singers from the Bavarian Radio Chorus (Chor des Bayerischen Rundfunks), all non-smokers and without pulmonic symptoms were included in the study (5 female, 5 male, age 44±11 years). All the subjects practice western classical singing and had completed their vocal studies at music conservatories. No subjects complained of dysphony: normal values were obtained for a German version of the singing voice handicap index 12 (SVHI-12)(*22*) (mean 2 ± 3).

### Tasks

All subjects were asked to perform the melody from the fourth movement of Ludwig van Beethoven’s 9th symphonyto the original text ("Freude schöner Götterfunken, Tochter aus Elysium”) written by Friedrich Schiller, in the key of D major, thus starting onF#3 for the male (fundamental frequency *f*_o_ approx. 185Hz) and F#4 (*f*_o_ approx. 370Hz) for female voices, respectively (Figure 1). This task is denoted as MT (melody and text). In a second task, the singers were asked to read the text aloud (task T) at a comfortable pitch, and in a third task to vocalize only the melody (task M) without text on the neutral vowel /ə/. All three tasks were performed with soft phonation (-) and loud phonation (+). Thus, 6 tasks were performed based on Beethoven’s 9^th^ symphony: MT+, MT-, M+, M-, T+, and T-. The sung tasks lasted approx. 6s corresponding to a speed of 80-90bpm for the MT and M task.

**Figure 1:**
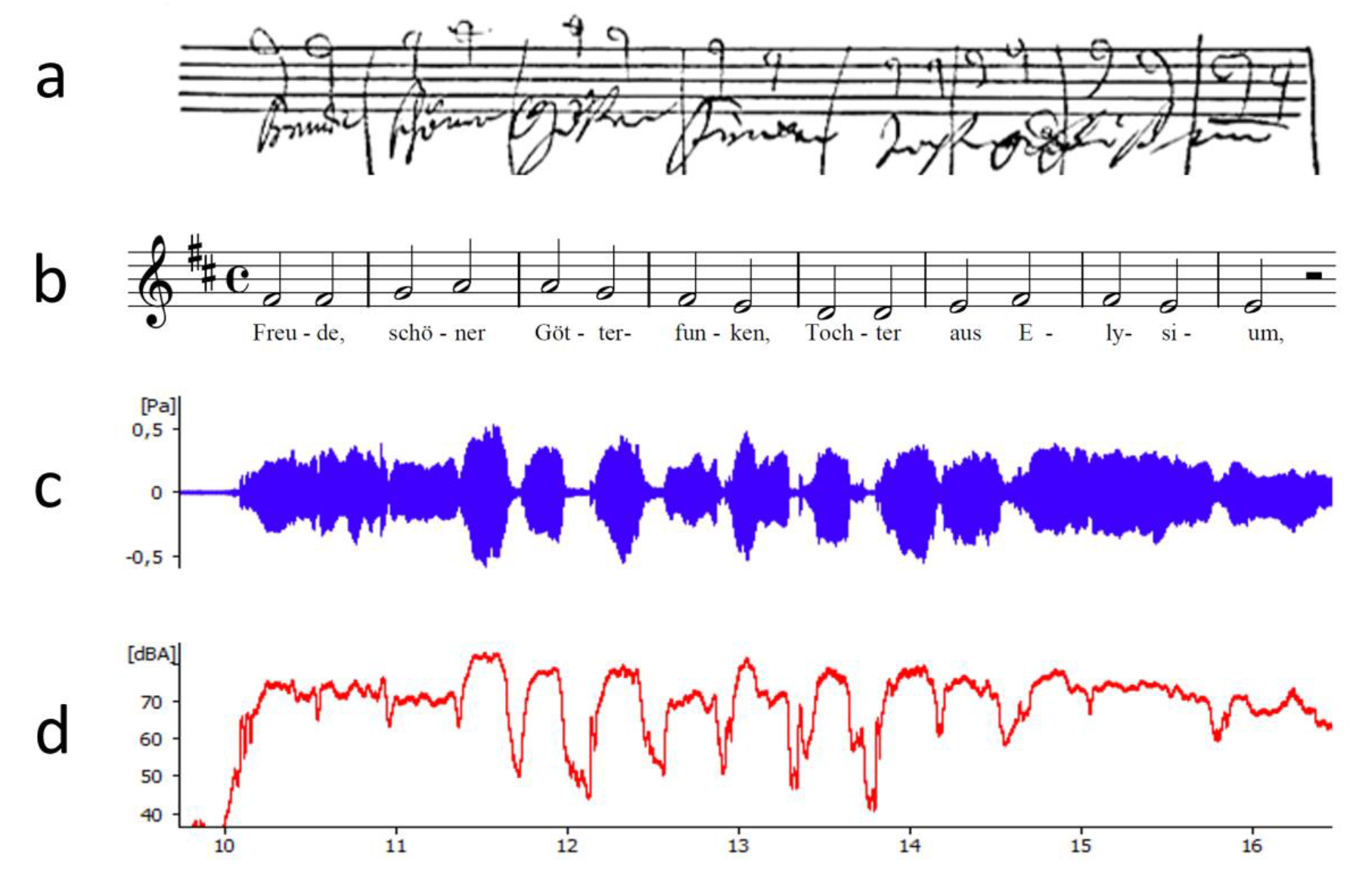
a) task in the original writing by Ludwig van Beethoven, b) task in the record, c) audio signal for MT+ subject 1, and d) sound pressure level for T+, subject 1.

For reference purposes, an exhalation over 6s and a coughing task were performed. In order to analyze the difference between consonants and vowels, in a single separate task, the subjects were asked to articulate consecutively the consonants /p/, /t/, /k/, /sch/ directly followed by the vowels /a/, /e/, /i/, /o/, /u/.

### Test setup

All measurements were performed in a Bavarian Broadcasting television network studio with the approximate dimensions: 27m × 22m × 9m (width × length × height). The setup is depicted in Figure 2. The singer stood on a lifting platform to lift him/her to the correct height. A mark at the singer’s forehead was used as a point of reference to adjust the height of the platform to the correct position, as shown in the zoomed parts of Figure 2. The wall was 4m behind and 5m to the left of the platform. To the right and in front of the platform the walls were more than 7m away. Three measuring rods for each spatial direction were mounted onto the lifting platform to convert the pixel dimension of the recorded picture into metric dimensions. The rods were assembled in a cross configuration and had a 2 cm and 10 cm scale, respectively.

**Figure 2:**
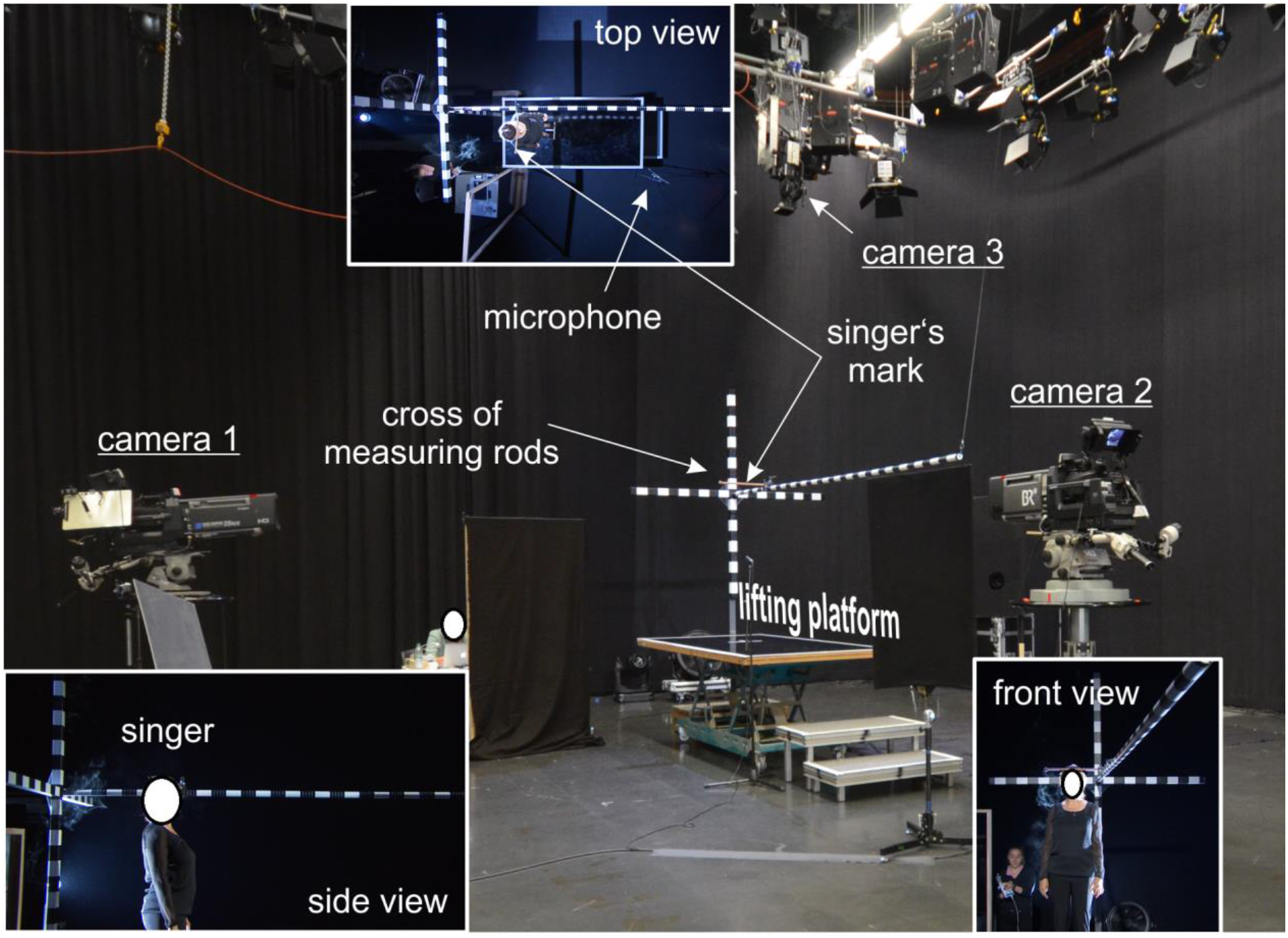
Picture of the test setup including the three cameras, the lifting platform and measuring rod cross (spatial directions are indicated by x, y and z). The perspective views of each camera are shown with a singer in performing position

Three full HD Sony HDC 1700R Bavarian Broadcasting cameras (resolution 1920 × 1080 pixels) recorded the singer and the cloud from a side view (camera 1), a front view (camera 2) and a top view (camera 3) perspective. The side and front camera were equipped with Canon DIGI SUPER 25 XS lenses (Canon, Tokio, Japan) and the top camera with a HD Fujinon HA14×4.5BERM/BERD wide angle HD lens (Fujinon, Tokio, Japan). All cameras recorded synchronously at a frame rate of 25 fps.

The audio signal was recorded with a Sennheiser KMR 81 directional and a ME62 omnidirectional microphone (Sennheiser electronic GmbH, Wedemark, Germany) which was placed at the lower right corner of the singer’s lifting platform, in ca. 1.5m distance to the mouth, as shown in the top view zoom in Figure 2. For estimation of the sound pressure level (SPL), the microphones were calibrated with the Sopran software (Svante Grandqvist, Karolinska Institut, Stockholm, Sweden) using a sound level meter (Voltcraft, Hong Kong, China).

For the experiments, all subjects inhaled the smoke of an e-cigarette filled only with the basic liquid which consists of 50% glycerin and 50% propylene glycol. To nebulize this liquid, a Lyneden Vox e-cigarette was used (Lynden GmbH, Berlin, Germany). According to Ingebrethsen et al.(*23*) the particles generated in e-cigarettes have a diameter in the range of aerosols at 250-450 nm. For each task, the singer’s individual inhaling volume was measured by a ZAN 100 spirometer (Inspire, Oberthulba, Germany) which was coupled with the mouthpiece of the e-cigarette. Being already on the platform, each singer was asked to inhale .5l smoke controlled by the spirometer. Immediately after inhalation, the singers moved to the singer’s mark and started to perform the task. The singers were instructed to maintain their position on the platform without moving for up to 60s after completing each task, to allow the exhaled cloud to be traced. Because all the singers were non-smokers, the inhalation was tested by the subjects before the experiments to become accustomed to the smoke. In rare cases of sudden coughing during the experiments, the task was repeated until it was performed correctly.

In order to achieve a large visual contrast between the cloud of smoke and the background, the entire studio was lined with black linen and the singers wore black clothes. The smoke was illuminated with three flash lights positioned on the left hand side of the platform: one in the left corner behind the platform with a distance of approx. 5m to the singer, one in front of the platform on the left side in a distance of 7m and another directly behind the platform approx. 3.5m from the rear edge of the platform.

Before each task, the studio was aerated with the main gate open for at least 2mins using a fan behind the platform. The main gate was at the opposite side of the studio with a distance of approx. 20m to the platform. Afterwards, the gate was closed and everyone present was instructed to stop moving for another 2 mins for air circulation that remained from the active aeration to settle down. Subsequently, the task started with the singer’s inhalation of the e-cigarette basic liquid. The temperature and the humidity in the studio were measured and recorded. The temperature was measured at mean 23.27°C (SD .46) and the relative humidity at 46.12% (SD .95), respectively.

For reference purposes, all tasks were performed a second time without inhalation of the basic liquid into a spirometer (ZAN100, Oberthulba, Germany) in order to check the real amount of exhaled air volume for such tasks.

### Video analysis

A coordinate system was defined to segment the video footage in three dimensions from the mouth of the singer (x-dimension to the front, y-dimension from the left to the right, z-dimension from the bottom to the top, Figure 3). To evaluate the temporal evolution of the exhaled smoke, the video footage of cameras 1 and 3 were converted into grayscale and the singer was covered with a black mask to avoid the segmentation of bright features such as skin or hair. The smoke cloud was then segmented in each video frame which was performed with an in-house software tool using a threshold-based region-growing algorithm yielding the area of the cloud and its contour as a function of time. The dimensions of the ROI are *ROIx* × *ROIy* × *ROIz* = 260*cm* × 270*cm* × 180*cm* for camera 1 and 190*cm* × 270*cm* × 180*cm* for camera 3. Based on the cloud contour, the maximum dimensions of the cloud (dx, dy, dz) as indicated in Figure 3 in each frame were computed. Thus, the dx and d_z_ dimension were calculated from camera 1 and d_y_ from camera 3.

**Figure 3:**
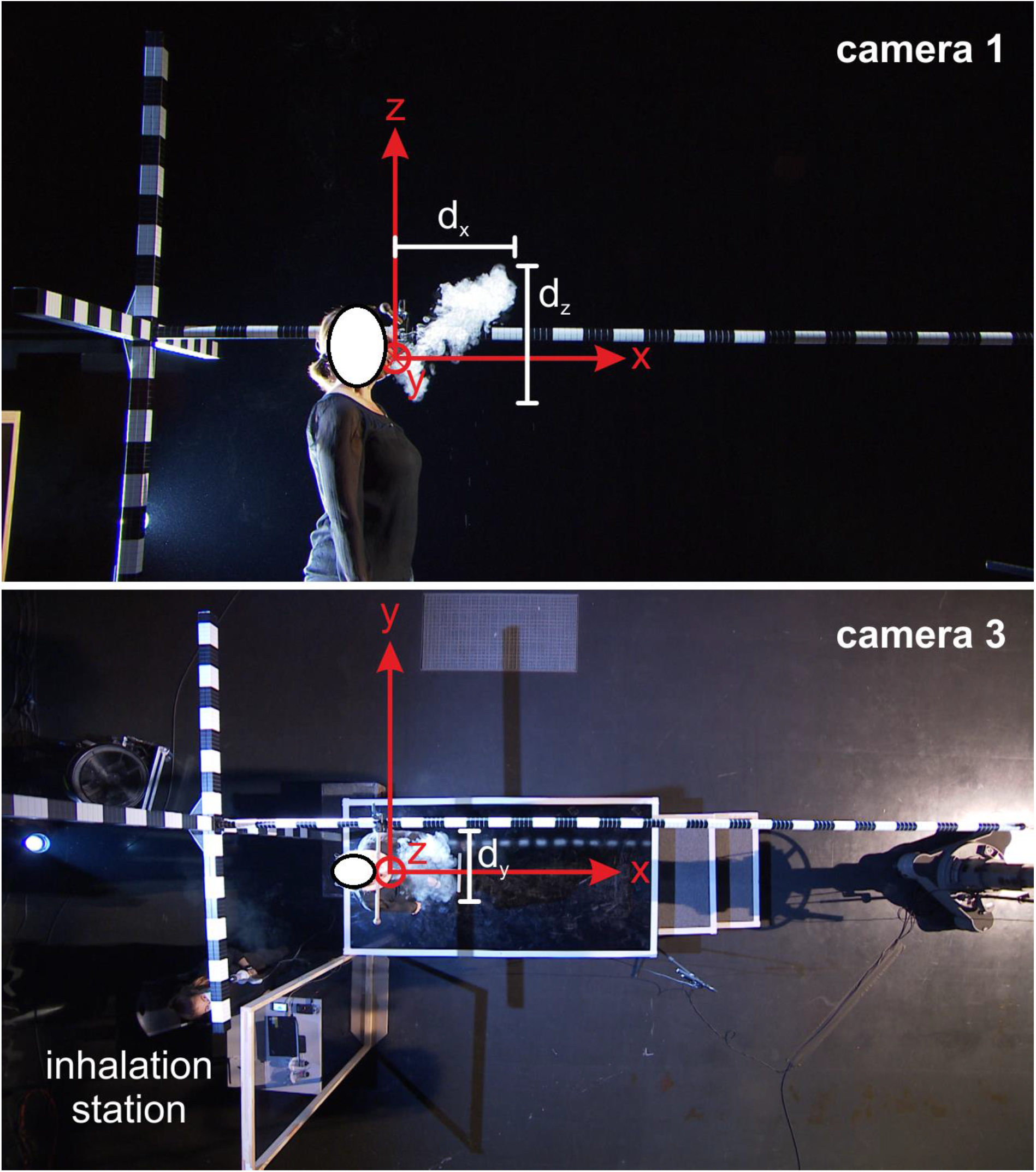
Pictures of camera 1 (top) and camera 3 (bottom) views with the defined coordinate system. Based on the segmentation of the cloud, its maximum dimensions dx, dy and dz were computed in each frame.

Figure 4 shows the dispersion of the cloud in each spatial direction. Several outliers occurred when non-smoke regions were segmented if the cloud reached a region with similar grayscale value as e.g. the white sectors of the measuring rods or remaining areas of bright skin of the singer’s face, neck and arms. Those outliers were removed in two steps: (1) a moving median filter with a fixed window length of 30 time points and (2) a subsequent cubic spline fitting approximation. The resulting curves are also shown in Figure 4 in black. The computation of curve smoothing was performed in Matlab (The Mathworks Inc., Natick, MA).

**Figure 4:**
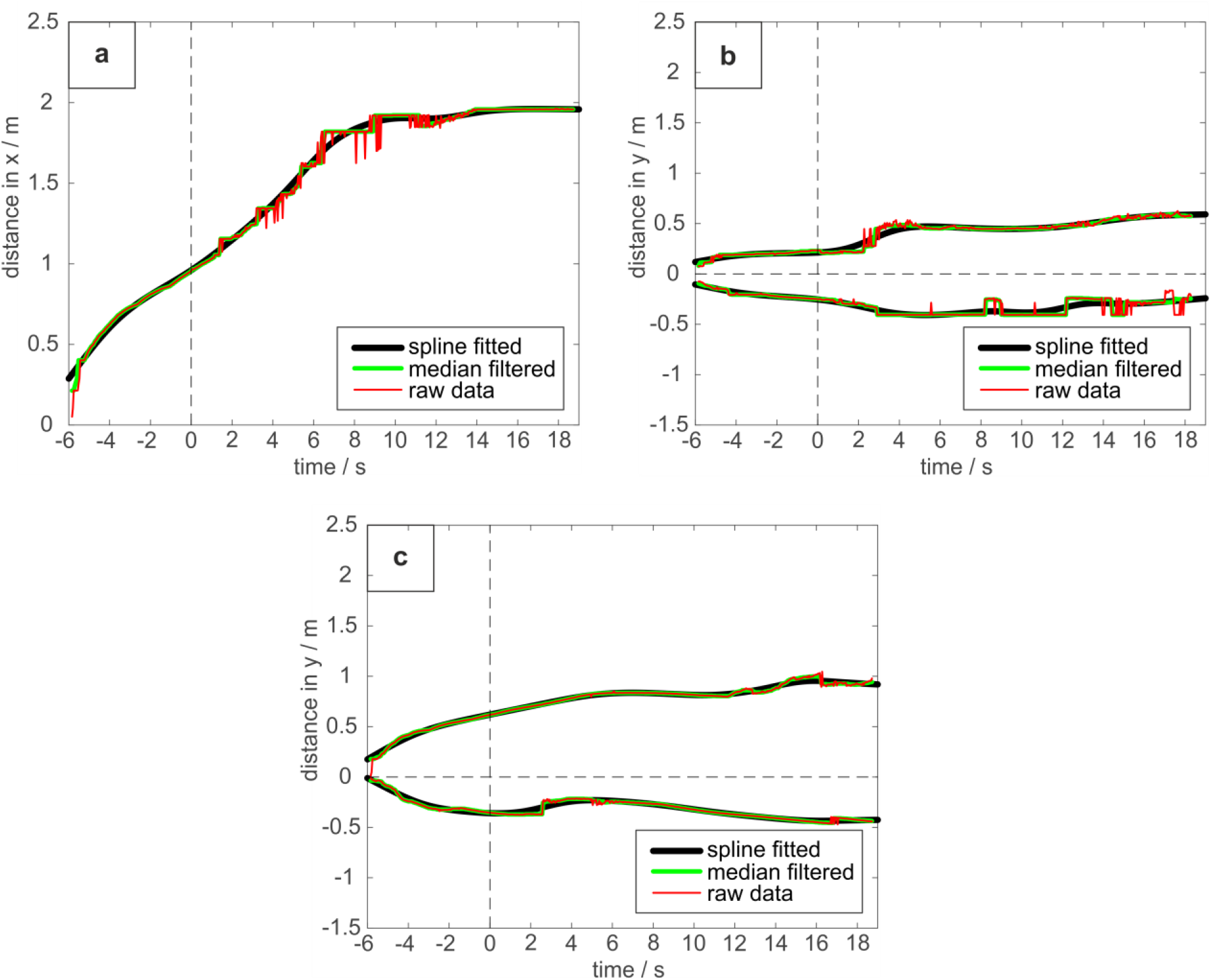
Expansion of the smoke cloud obtained from the raw segmentation (red), after median filtering (green) and spline fitting (black) as function of time. The distance from the mouth (x,y,z=0) is shown for the x-direction in subfigure (a), for the y-direction (left-right diameter) in (b) and for the z-direction (upward-downward diameter) in (c).

### Statistics

Because of the small number of subjects no comparative statistical analysis appeared meaningful. Instead, all traces for all subjects are provided in the supplementary material.

## Results

In the experiment without inhalation of the basic e-cigarette liquid, the subjects exhaled a comparable air volume for all tasks. For the experiment, the inhaled volume of basic liquid was comparable among the tasks as displayed in table 1.

**Table 1:** Exhaled volumes measured for all task before the experiment without inhalation of basic liquid, inhaled volume of basic liquids and measured sound pressure level (SPL).

| task | Exhalation without smoke (ml) |  | Inhaled Volume basic liquid (ml) |  | SPL (dB(A)) |  |
| --- | --- | --- | --- | --- | --- | --- |
|  | mean | SD | mean | SD | mean | SD |
| MT- | 1787 | 661 | 476 | 162 | 57.08 | 3.38 |
| MT+ | 1958 | 679 | 507 | 230 | 67.75 | 4.24 |
| T- | 1654 | 490 | 494 | 252 | 44.69 | 4.40 |
| T+ | 1675 | 512 | 511 | 215 | 65.32 | 6.84 |
| M- | 1726 | 520 | 493 | 215 | 61.74 | 6.02 |
| M+ | 1977 | 830 | 592 | 222 | 73.12 | 4.69 |

With regard to the impulse dispersion measured at the end of each task, the mean and median distances across all subjects are displayed in table 2 for all three spatial directions (x,y,z) and Figure 5 shows the representative images for the point of completion of all 6 tasks performed by a female subject (the corresponding videos can be found in the supplementary data). Considering the dispersion to the front (x-direction), the mean distance from the mouth was .85m for MT+ and .83m for MT-measured at the end of both MT tasks. Accordingly, the distances for T+ and T-were only slightly lower with .79m (T+) and .75m (T-). In contrast, M+ and M-revealed distinctively lower values with .63m and .56m for M+ and M-, respectively. Although the median expansion of the aerosol cloud in all directions was below 1m at the end of the task, the inter-subject variability was large ranging from .61m up to 1.36m for the MT tasks in x-direction. Figure 6 represents the traces for all subjects and all directions for task MT+ and Figure 7 shows the median values for all tasks and directions. The traces for the other tasks for all subjects are provided in the supplementary data. As is also evident in these figures, the expansion of the aerosol cloud in the the y- and z-direction was smaller than in the x-direction except for the M± tasks, which exhibit similar expansion of the aerosol cloud to either side. Furthermore, the distances in y-direction show for some subjects an imbalance between the left and right side of expansion, presumably due to a small convectional flow generated by the singer’s motion. This imbalance can be seen in Figure 6b and 7b. Here, the cloud expansions to the left (positive y range) do not show a symmetrical expansion to the right side (negative y range) of a subject. In order to compensate for the convectional flow, figures 6d and 7d show the y-diameter from the left to the right exhibiting median values between .57m and .87m at the end of the task.

**Table 2:**
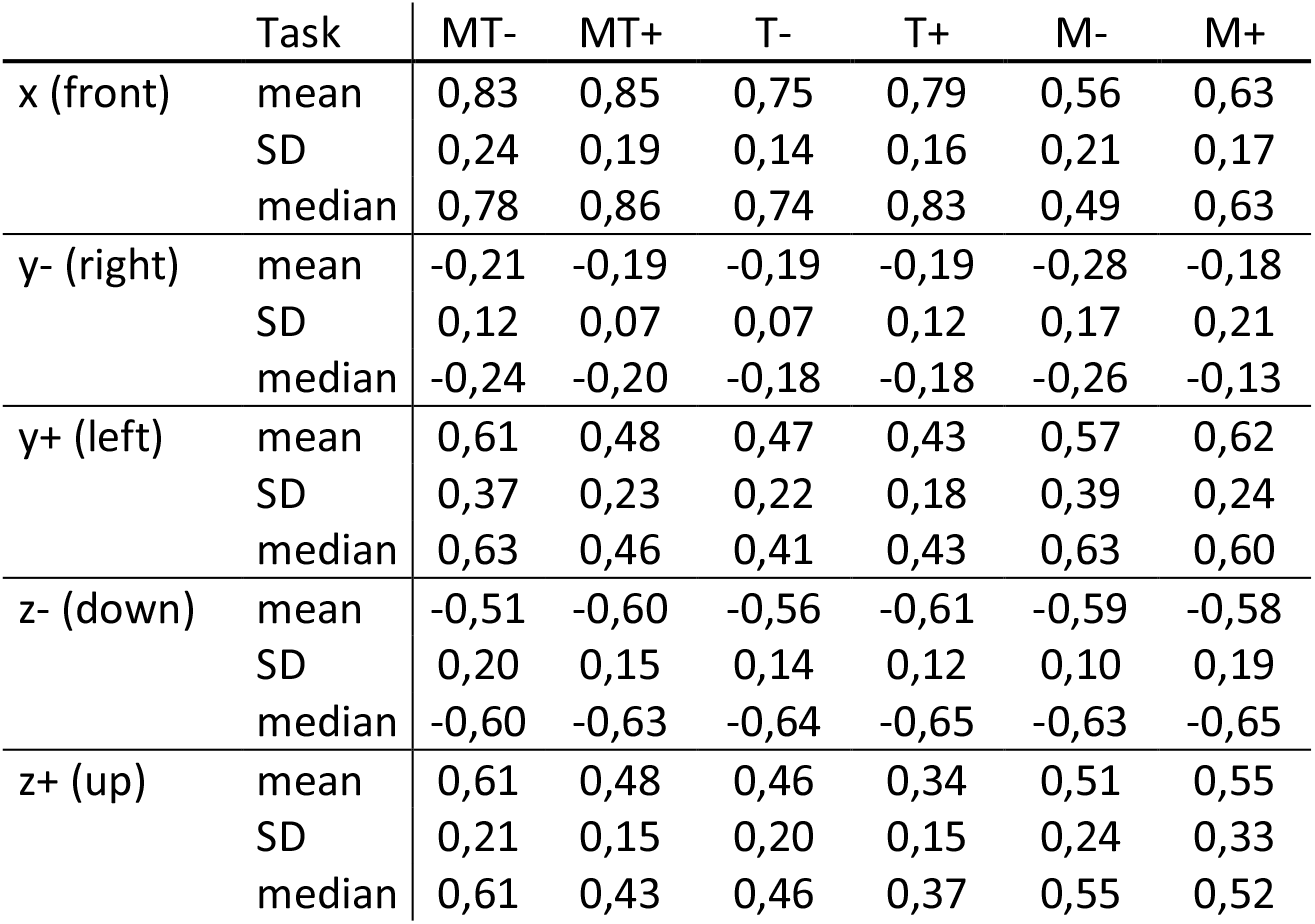
Mean values, standard deviation (SD) and median at the end of the task for all three physical directions directions of propagation.

| Task |  | MT- | MT+ | T- | T+ | M- | M+ |
| --- | --- | --- | --- | --- | --- | --- | --- |
| x (front) | mean | 0,83 | 0,85 | 0,75 | 0,79 | 0,56 | 0,63 |
|  | SD | 0,24 | 0,19 | 0,14 | 0,16 | 0,21 | 0,17 |
|  | median | 0,78 | 0,86 | 0,74 | 0,83 | 0,49 | 0,63 |
| y- (right) | mean | -0,21 | -0,19 | -0,19 | -0,19 | -0,28 | -0,18 |
|  | SD | 0,12 | 0,07 | 0,07 | 0,12 | 0,17 | 0,21 |
|  | median | -0,24 | -0,20 | -0,18 | -0,18 | -0,26 | -0,13 |
| y+ (left) | mean | 0,61 | 0,48 | 0,47 | 0,43 | 0,57 | 0,62 |
|  | SD | 0,37 | 0,23 | 0,22 | 0,18 | 0,39 | 0,24 |
|  | median | 0,63 | 0,46 | 0,41 | 0,43 | 0,63 | 0,60 |
| z- (down) | mean | -0,51 | -0,60 | -0,56 | -0,61 | -0,59 | -0,58 |
|  | SD | 0,20 | 0,15 | 0,14 | 0,12 | 0,10 | 0,19 |
|  | median | -0,60 | -0,63 | -0,64 | -0,65 | -0,63 | -0,65 |
| z+ (up) | mean | 0,61 | 0,48 | 0,46 | 0,34 | 0,51 | 0,55 |
|  | SD | 0,21 | 0,15 | 0,20 | 0,15 | 0,24 | 0,33 |
|  | median | 0,61 | 0,43 | 0,46 | 0,37 | 0,55 | 0,52 |

**Figure 5:**
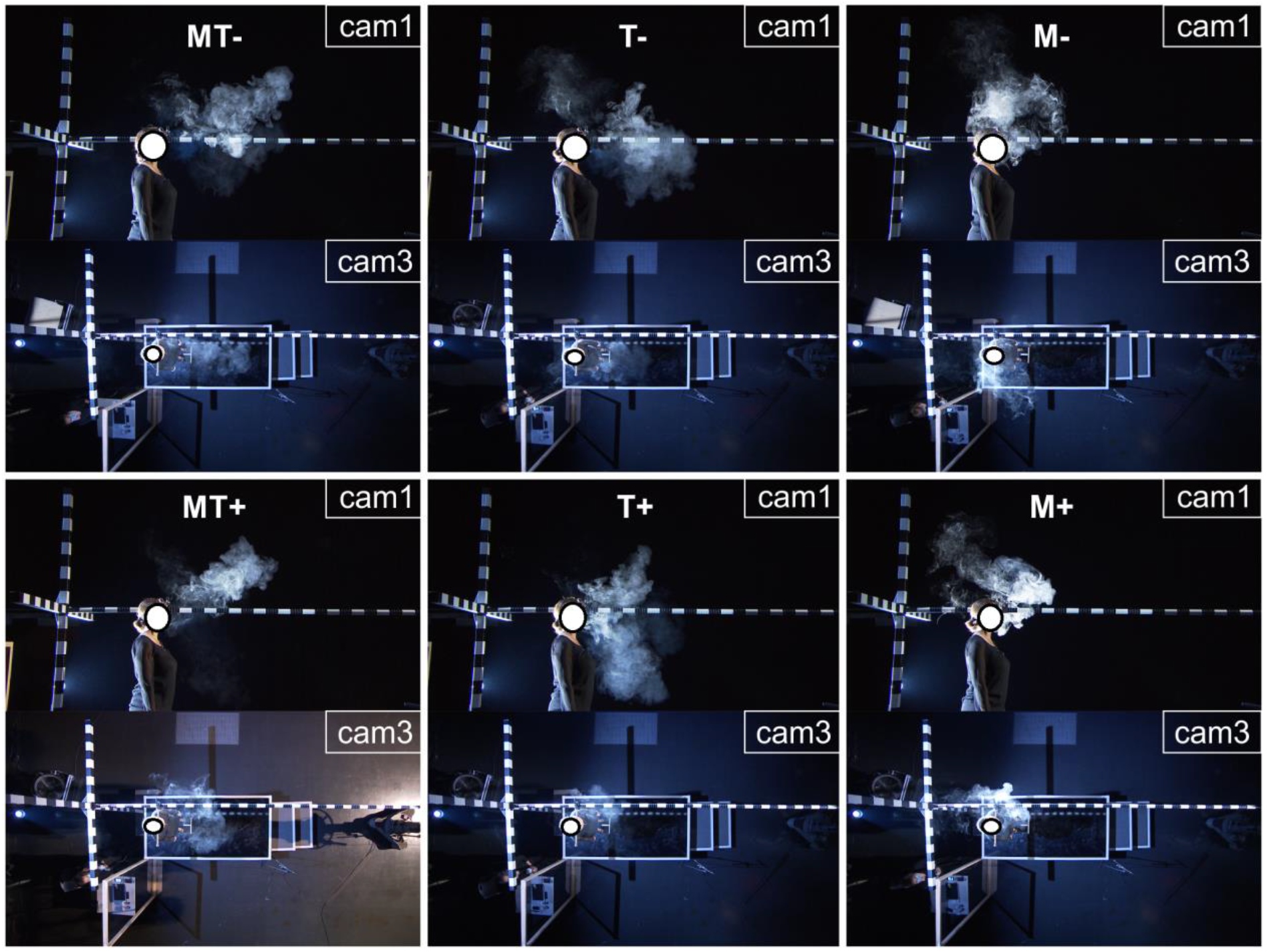
Pictures of camera 1 (top) and camera 3 (bottom) views for the end of a task. The tasks MT±, T± and M± are shown from left to right, the soft tasks in the top and the loud tasks in the bottom line.

**Figure 6:**
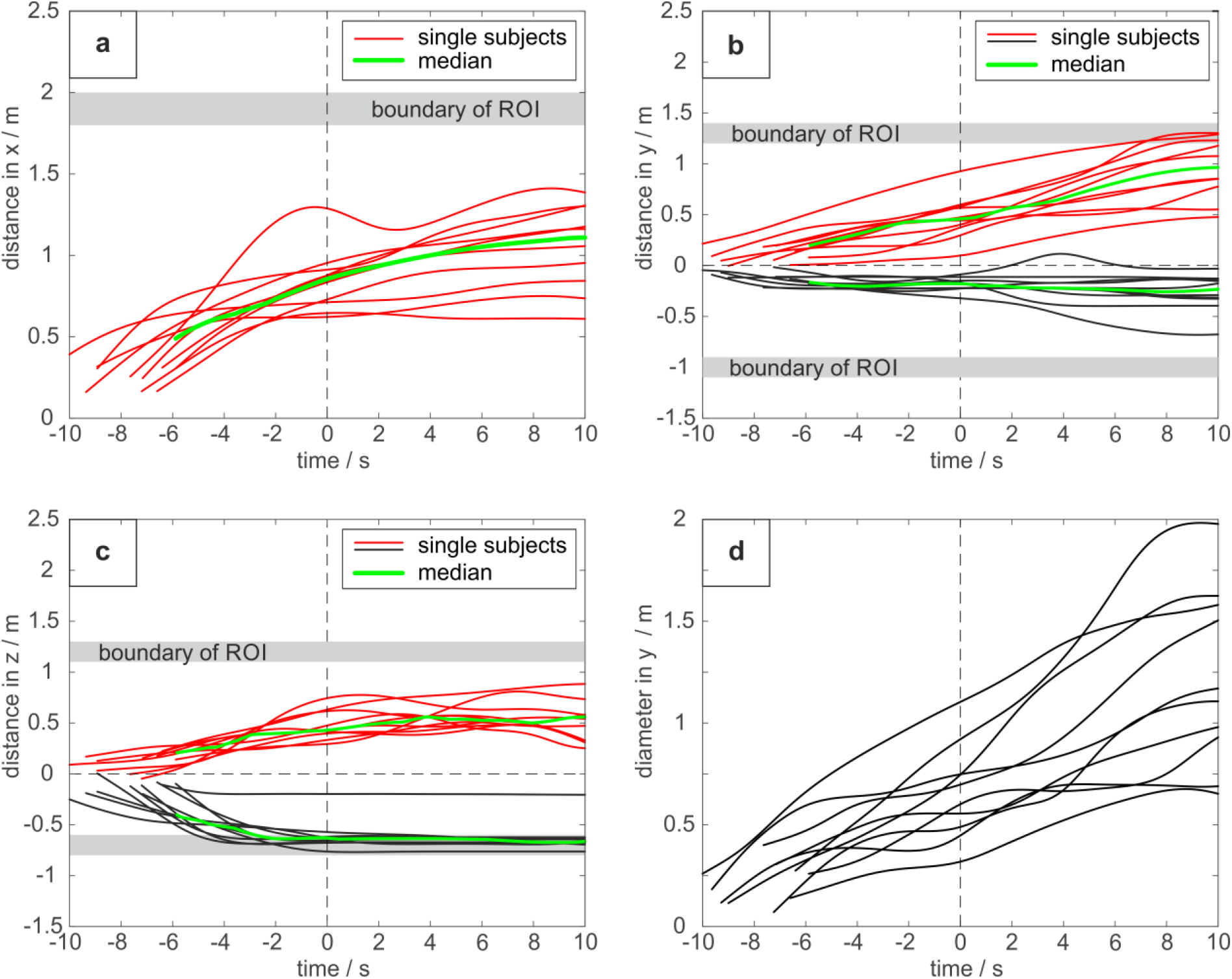
Traces of all subjects for all dimensions (a: x-dimension (front), b: y-dimension (left-right), c: z-dimension (Up-down) and d: diameter of the y-dimension) for the MT+ task. The 0 point in the time-scale refers to the end of the task. The green lines show the median. The corresponding graphs for the other tasks are provided in the supplemental material.

**Figure 7:**
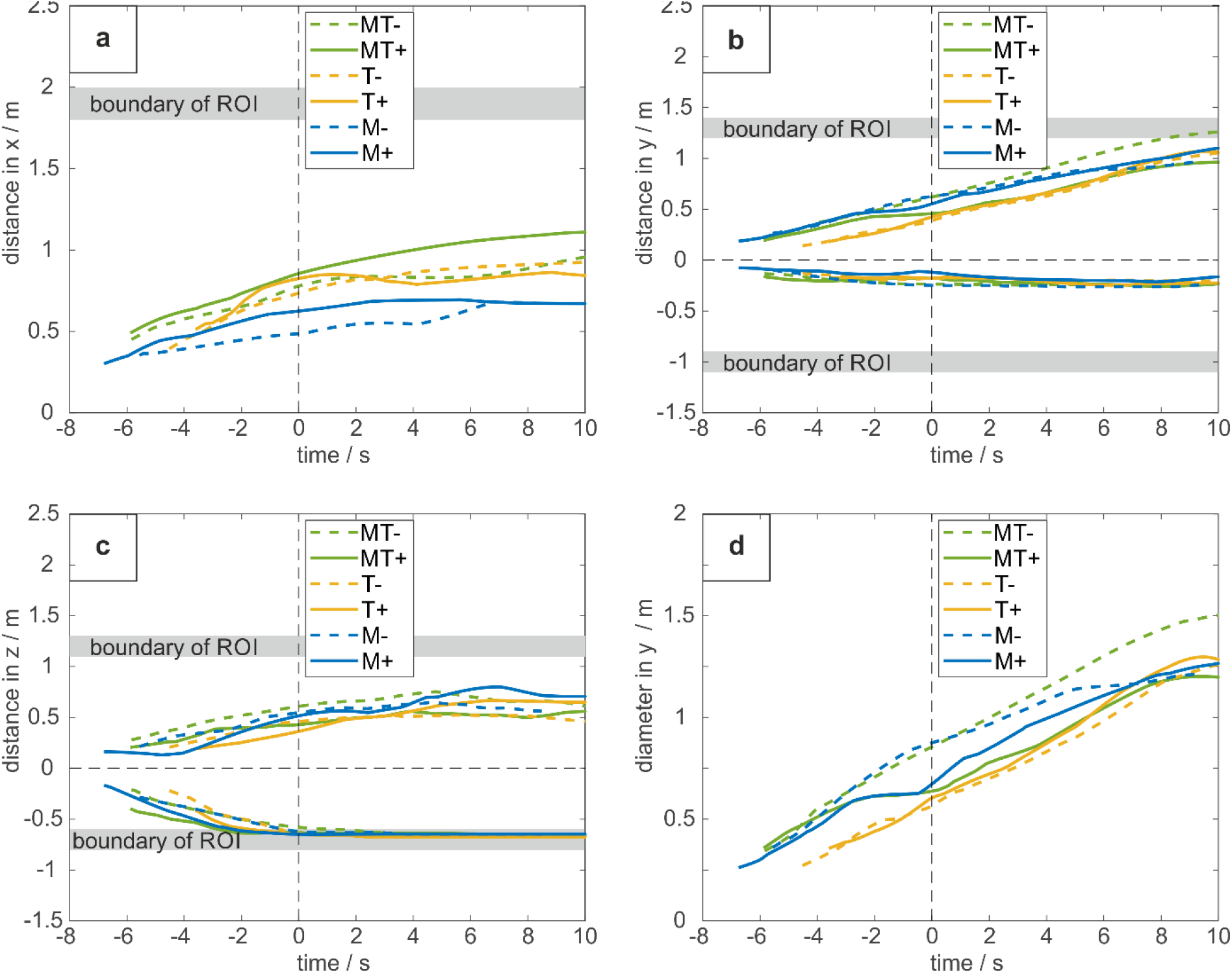
Median-traces for all dimensions (a: x-dimension (front), b: y-dimension (left-right), c: z-dimension (Up-down) and d: diameter of the y-dimension) and all tasks. The 0 point in the time-scale refers to the end of the task. The green lines show the median. The corresponding graphs for the other tasks are provided in the supplemental material.

Once a task was complete the motion of the aerosol cloud decreased. To the front, the additional movement was on average between .04m and .11m for all tasks 3s after the end of task. The movement to the left was greater reaching additional distances between .14 and .21m owing to the small convectional flow mentioned above. In contrast, to the right hand side, there was nearly no additional movement (0 to .02m) 3s after the task. The movement upwards was greater reaching additional distances of .05 to .15m mean difference between the end of the task and 3s after its completion.

Considering the aerosol dispersion for soft and loud tasks, the singing tasks M and MT produced a larger distance for loud singing in comparison to their soft equivalent as shown in Figure 7a. For the T task, this difference was not evident. In contrast to the x-direction, the aerosols moved farther in positive y- and z-direction during the soft tasks M- and MT-as shown in Figure 7b and 7c. Again, there was no large difference between T- and T+ in y- and z-direction. Accordingly, these trends also hold for the y-diameter of the aerosol cloud shown in Figure 7d: there were larger diameters for the soft tasks M- and MT- and no clear difference between T- and T+.

It could be assumed that the smaller distances for M± arise from the absence of consonant articulation compared the MT± and T± tasks which produce a large impulse owing to their plosive and fricative characteristics. To evaluate the difference in the aerosol dispersion to the front between consonant and vowel production,, the separate task with the consecutive articulation of the consonants /p,t,k,sch/ followed by the vowels /a,e,i,o,u/ articulation is shown in Figure 8 for the x-direction. The asterisks represent the time point .2s before the /a/ articulation started. Thus, the dispersion left of the asterisks represents the distance reached by the consonants. Except for the subject represented by the topmost curve, the greatest increase in x-distance is produced during the consonant articulation. During the subsequent phonation of vowels, however, the distance increased less and in some cases the cloud didn’t extend any further in the x-direction.

**Figure 8:**
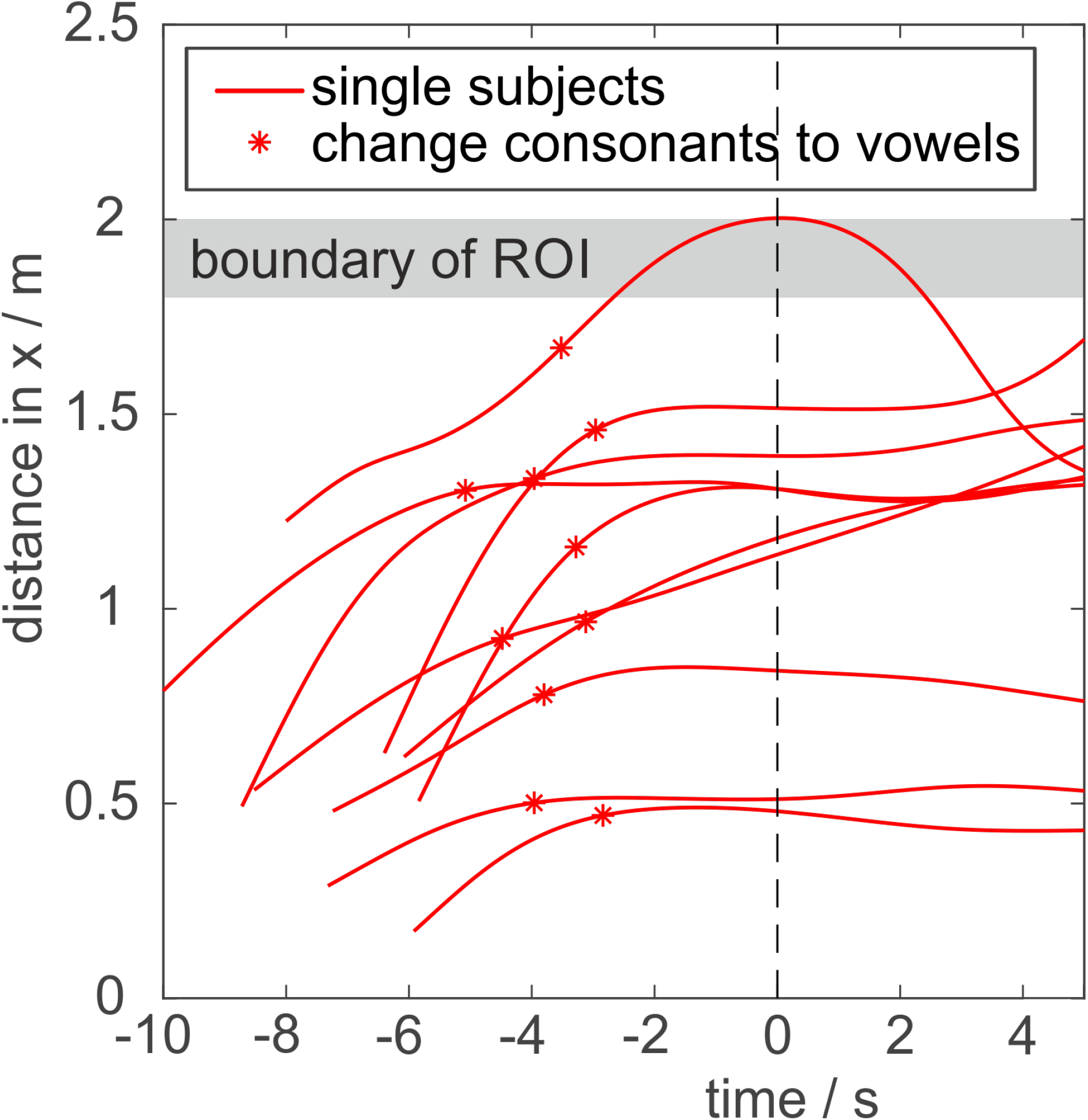
Aerosol cloud dispersion in x-direction for the additional task of consonants /p,t,k,sch/ and subsequent vowels /a,e,i,o,u/ for all 10 subjects. The time point 0s marks the end of the task. The asterisks represent the time points .2s before the particular subject started the vowel phonation.

With regard to both breathing and coughing, detected distances were much greater than all phonation related tasks. After 6s exhalation, the mean distance in the x-direction averaged 1.19m with a maximum of 1.71m. Similarly, after coughing, the mean distance of 1.32m with the maximum at 1.89m was observed.

## Discussion

Aerosols are assumed to contribute to person-to-person transmission of CoVID19(*11-13*). The presented data showed that the impulse spreading of aerosols from singing a text reach distances to the front of up to 1.4m in a short amount of time. Furthermore, the data reveal that the distance of dispersion of singing a text is comparable to reading a text rather than singing only on a single vowel.

In order to protect other people from potentially infectious particles during the CoVID19 pandemic, it seems important to understand the impulse dispersion dynamics for aerosols. With respect to the expulsion to the front, the added basic liquid aerosols reached mean distances of more than .8m from the mouth on completion of the MT+ and MT-task. However, many subjects reached distances greater than 1m and some subjects even reached 1.4m. Still, these distances were lower than the distances reached through a 6s exhalation. In agreement with Bourouiba(*4*) coughing also revealed much greater distances in the presented study, with a mean of 1.3m and maximum of 1.9m. In contrast to singing a text or speaking, singing a vowel exhibited much lower values for both tasks, soft (M-) and loud (M+). Because such M- or M+ tasks are somewhat artificial in singing practice, it seems reasonable to formulate recommendations for distances between choir singers based on the impulse dispersion of aerosols for the MT tasks rather than M. As a consequence, distances of 1.5m between persons in the frontal direction which are sometimes recommended for protection during choir rehearsals or at religious services(*24*) should be increased to a distance of at least 2m(*8*) which could be estimated borderline.

The dispersion distance to the side was found to be much lower. In order to avoid any convection flow in the room, the flash lights and cameras were positioned far away from the singer, who was about 5m from the nearest wall, the flash lights were covered and the fan was turned off. However, there was a slight convectional flow to the left for some subjects. The reason remains unclear. However, a reasonable explanation for the production of the small convectional flow is the movement of the subject just before the task started. The singers inhaled the aerosols at the right rear corner of the platform as shown in the camera 3 perspective in Figure 2. After the inhalation, the subject rotated counterclockwise and stepped forward to the singer’s mark potentially inducing a small convection flow to the left side. An example of this can be seen in the camera 3 perspective for task M+ shown in Figure 5. Given that the entire aerosol cloud is shifted to the left side, the diameter in the y-direction was evaluated in order to estimate the distance to one side without this convection flow. This measurement can then be regarded the distance either side in transverse (y) direction which is much lower than the distances to the front (x-direction). Taking these factors into account, it could be assumed that the chosen protective distance to the side could be lower than the distance to the front. The mean elevation distance of the aerosols reached .6m to the bottom and .5m to the top for MT+ at the end of the task. Due to the relatively minor role this dimension usually plays in the person-to-person distance of a choir, it is considered of less relevance for safety distance recommendations.

After the impulse dispersion the aerosols remain in the air and continue to move, although slowed down as they are mainly driven by their inertia. The aerosols only reached an additional distance of .11m to the front and .21m to the side three seconds after the tasks were complete. Thus, the distance reached by the initial impulse was much greater and, consequently, are considered more relevant for estimating safety distances for singers in order to reduce direct person-to-person transmission of potentially infectious particles. However, it could be assumed that inhalation from a virus accumulated cloud of aerosols could also play a major role in the virus transmission(*25*) if the aerosols are not removed from the air. Hence, it appears meaningful to investigate how much aerosols accumulate over time with regard to phonation dose and size of the room. Moreover, such results could contribute to recommendations for how to remove aerosols from the room, for example by aeration of fresh air. Also in this context, the presented data show only dispersion dynamics of the aerosol producer. Another aspect is whether the risk of person-to-person transmission during singing is increased due to the special deep breathing patterns of the performing singers.

The presented study used an artificially added aerosol with a comparable amount of aerosols for each task. The realistic number of aerosols expelled during singing is, however, much lower. It has been found that for voiced counting the number of expelled droplets with sizes of .3 ≤ D ≤ 20µm was .322 cm^-3^. Moreover, during singing the amount of expelled particles of these sizes was 1.088cm^-3^. Therefore particle emission rates are ten times higher during singing than during breathing and three times higher than during voiced counting(*17*). Furthermore, during speaking the amount is lower for a greater rate of consonants and higher for a greater rate of vowels within the spoken passage. In a recent pre-print Mürbe et al. estimated that singing with text produces up to 100 times more particles than speaking and up to 330 times more particles than beathing(*19*). However, the text differed between the singing and speaking task. To the best of the authors’ knowledge, there is no data available analyzing whether the absolute amount of aerosol emissions whilst singing a text is also greater than speaking exactly the same text passage. However, since vowels are phoned over a longer period of time during singing in comparison to speaking, also duration of the vocal fold oscillations, which has been assumed to be the reason for the increase of aerosols(*26*), is greater. Consequently, the risk of person-to-person transmission by aerosols is estimated to be greater for tasks with longer passages of vowel phonation and less consonant articulation. However, the present study observed the dispersion distance to the front to be much lower for the corresponding M± tasks, reducing the transmission risk for the impulse. This was also confirmed by the task with the separate consonant and vowel articulation which featured the greatest dispersion distances by the consonants in comparison to a much smaller and sometimes no distance increase by the vowel phonation. Thus, the lower distances for the M± tasks is most likely caused by the lack of consonants.

The softer tasks showed slightly lower dispersion to the front than the louder tasks, particularly for MT and M. Loudness and the sound pressure level are dependent on the pressure difference across the vocal folds during phonation(*27*). The greater pressure differences should contribute to greater air flow and therefore greater dispersion of aerosols to the front. In this respect, the largest frontal dispersion was in agreement with Bourouiba et al., who found similar for coughing produced by the sudden release of a large subglottal pressure below the vocal folds(*4, 26*). Furthermore, Asadi et al. found that the absolute aerosol production was greater for louder phonation(*2*). As a consequence the potential transmission risk appears increased for loud phonation resulting from both the absolute aerosol production and the greater dispersion distance to the front.

## Limitations

The presented study analyzed the standard text of “Freude schöner Götterfunken, Tochter aus Elysium”. It cannot be excluded that different texts or different languages would show different results. However, Schiller’s text with Beethoven’s melody has the great advantage that both, the melody and the text are well known for professional singers minimizing artifacts due to learning effects. Furthermore, the text offers multiple plosive and fricative consonants demonstrating a typical sample of sung and spoken text.

Also in this context, the presented study addresses the aerosol dispersion during a phonation time of about 6 to 8 seconds for all tasks. Therefore, the phonation time was too long to study all vowels and consonants used during the text in detail. That was the reason to include distance measurements for some consonants and vowels in a separate recording for comparison. However, the phonation time of the main tasks was much shorter than a realistic singing dose in rehearsals or concerts. It could be expected that the accumulation of aerosols in a closed room would be dependent on the phonation dose. Because the impulse dispersion stopped after a short amount of time, it could be assumed, however, that a longer phonation time would not greatly affect the measured distances for the aerosol impulse dispersion.

Owing to the large amount aerosols, the inhaled gas mixture possesses a higher density. This changed some subjects’ self-awareness during singing – presumably due to changes of the resonance properties of the vocal tract. Because only professional singers were included, these subjects could, nevertheless, easily compensate for these changes during the experiment, able to apply their singing technique under the experimental circumstances. Moreover, the fact that only professional singers were included could have influenced the articulation patterns for consonants. Because singers are trained to overexpress consonants for better text intelligibility during singing, the distances could be greater than for non-professional singers. It would be important to analyze if the dispersion distance from the mouth is lower for amateur singers, whereby closer proximity with protection from impulse spreading for both droplets and aerosols might be safe.

Last, because only professional singers were included the number of subjects was too small to perform comparative statistical analyses. It is be hoped that a large number of singers will be included in future studies in order to test the observed differences.

## Conclusions

This study analysed the impulse dispersion of aerosols in professional singers. The impulse dispersion to the front was found to be greater than in the transverse direction to the sides. For safety distances 1.5m to the front and 1m to the side appear too short. Consequently, a distance of at least 2-2.5m to the front and 1.5m to the side should be recommended for choir rehearsals or singing at religious services. However, safety is also dependent on accumulation of aerosols during phonation and convectional flow. Therefore, a continuous aeration during phonation could diminish the amount of aerosols and therefore reduce the risk of infection transmissions. Thereby, it is important that the forced aerating flow is not guided through other attendant persons in the room.

## Data Availability

not yet

## Acknowledgements

The authors thank all members of the Bavarian Broadcast for their help in realizing this study. Also, the authors thank Donata Gellrich, MD, for help in the design of the study and Helena Daffern, PhD, for native corrections.

## Contribiutions

**ME:** Study Design, Performing experiments, Analysis, Manuscript draft

**SG:** Performing experiments, Analysis, Manuscript draft

**GP:** Performing experiments, Analysis, Manuscript draft

**CW:** Performing experiments, Analysis, Manuscript draft

**TB:** Study Design, Performing experiments, Manuscript draft

**BJ:** Performing experiments, Manuscript draft

**LK:** Analysis, Manuscript draft

**MD:** Analysis, Manuscript draft

**SK:** Study Design, Performing experiments, Analysis, Manuscript draft

---

**Supplemental Figures 6:**
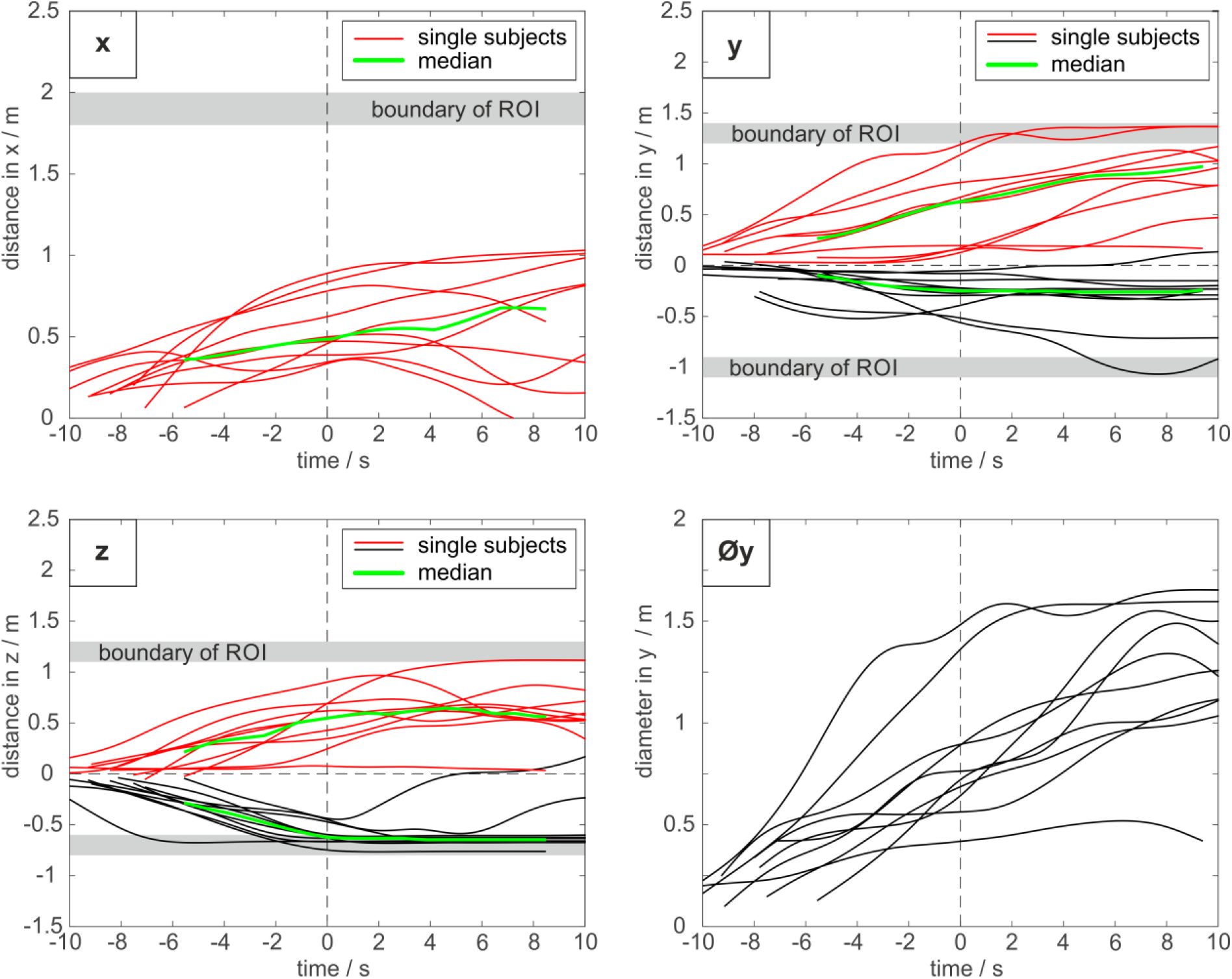
Traces of all subjects for all dimensions (a: x-dimension (front), b: y-dimension (left-right), c: z-dimension (Up-down) and d: diameter of the y-dimension) for the M-task. The 0 point in the time-scale refers to the end of the task. The green lines show the median. The corresponding graphs for the other tasks are provided in the supplemental material.

**Supplemental Figures 6:**
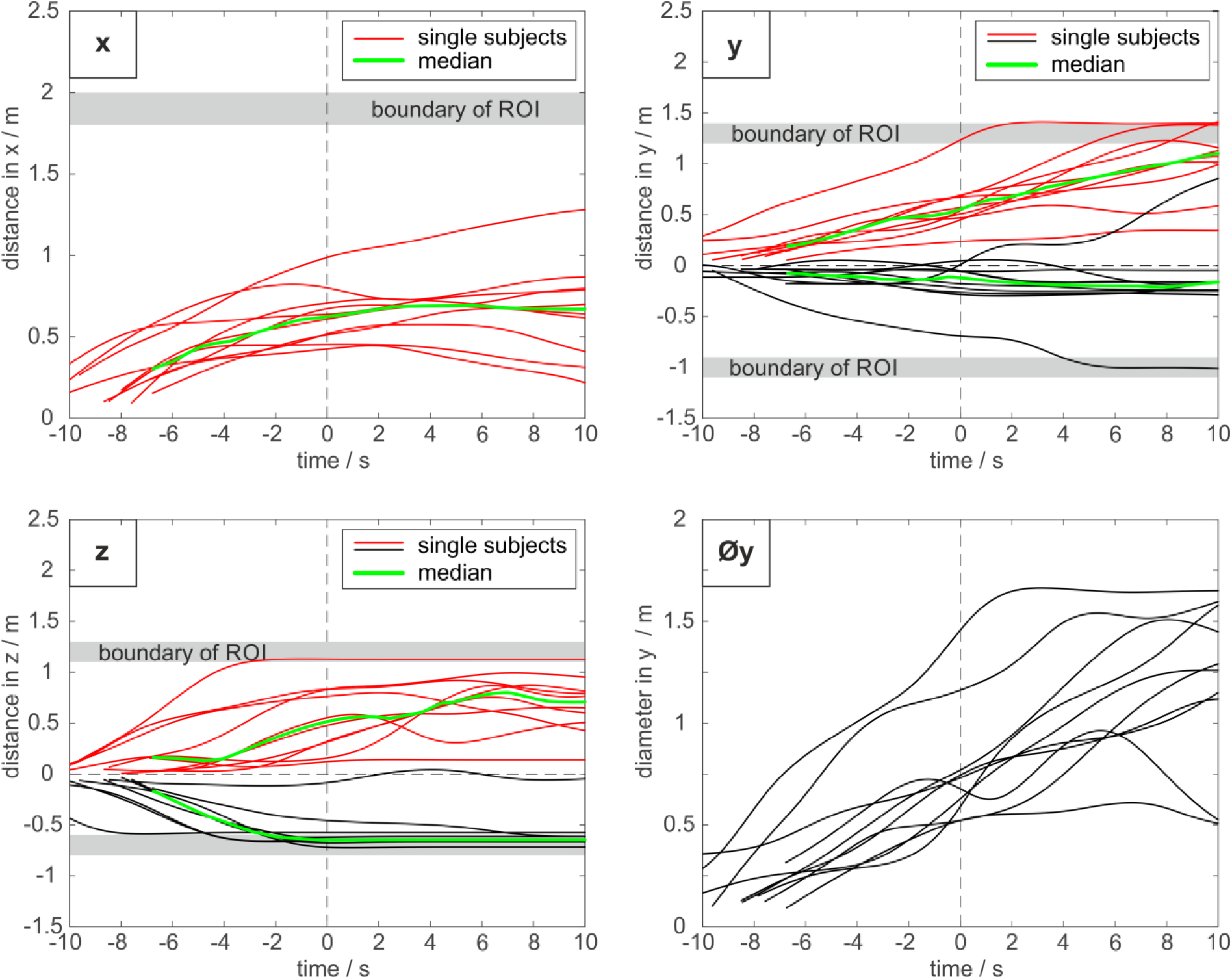
Traces of all subjects for all dimensions (a: x-dimension (front), b: y-dimension (left-right), c: z-dimension (Up-down) and d: diameter of the y-dimension) for the M+task. The 0 point in the time-scale refers to the end of the task. The green lines show the median. The corresponding graphs for the other tasks are provided in the supplemental material.

**Supplemental Figures 6:**
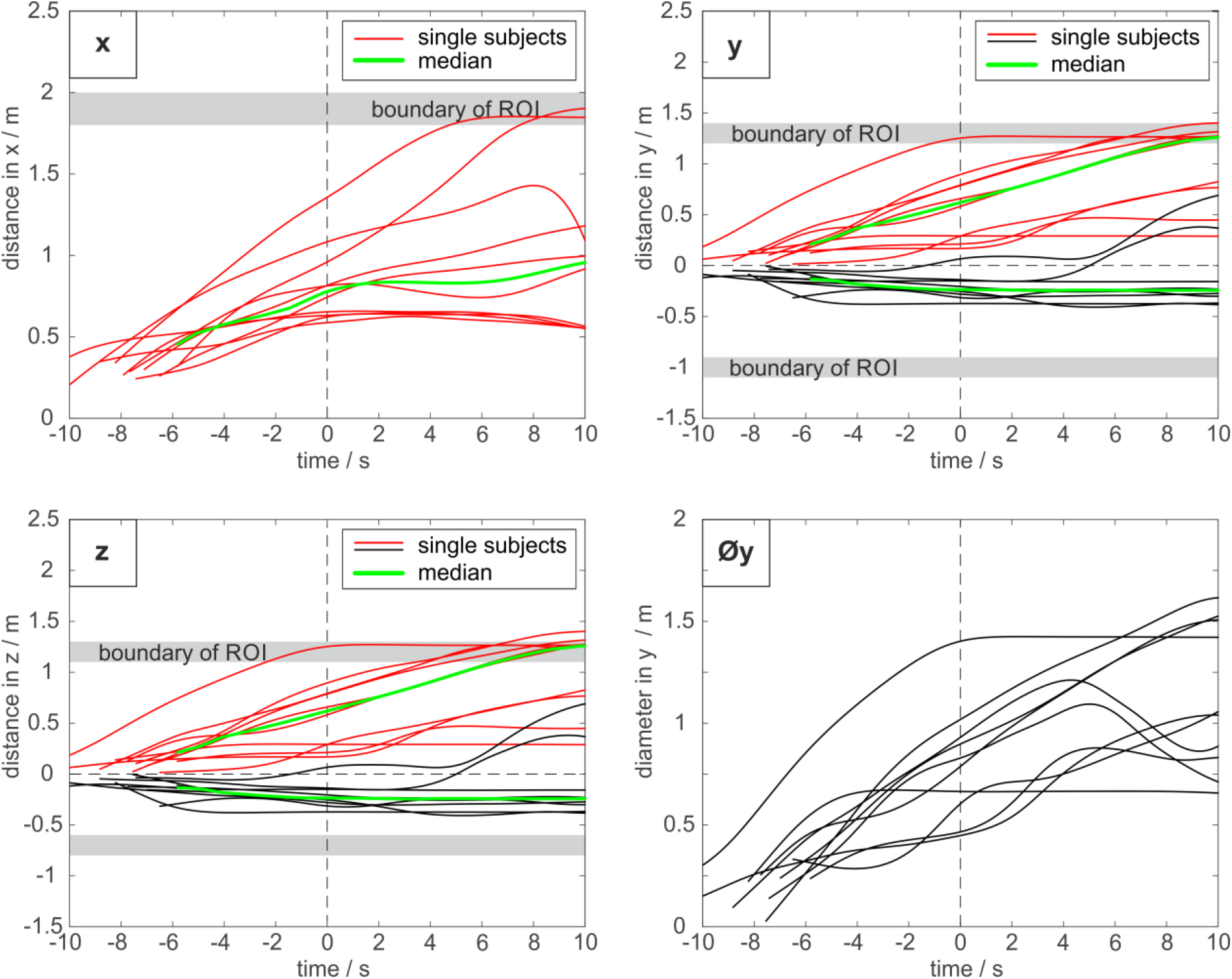
Traces of all subjects for all dimensions (a: x-dimension (front), b: y-dimension (left-right), c: z-dimension (Up-down) and d: diameter of the y-dimension) for the MT-task. The 0 point in the time-scale refers to the end of the task. The green lines show the median. The corresponding graphs for the other tasks are provided in the supplemental material.

**Supplemental Figures 6:**
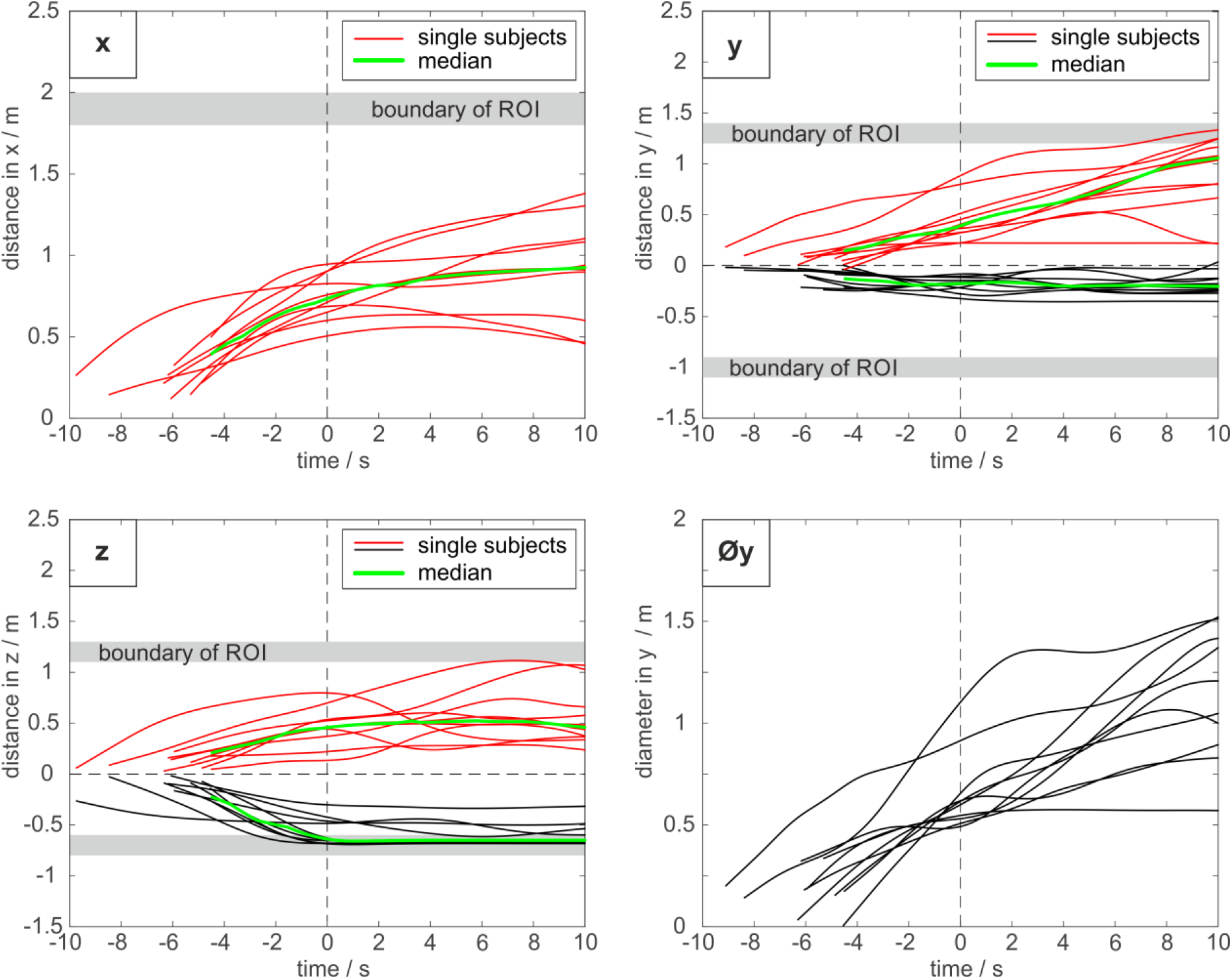
Traces of all subjects for all dimensions (a: x-dimension (front), b: y-dimension (left-right), c: z-dimension (Up-down) and d: diameter of the y-dimension) for the T-task. The 0 point in the time-scale refers to the end of the task. The green lines show the median. The corresponding graphs for the other tasks are provided in the supplemental material.

**Supplemental Figures 6:**
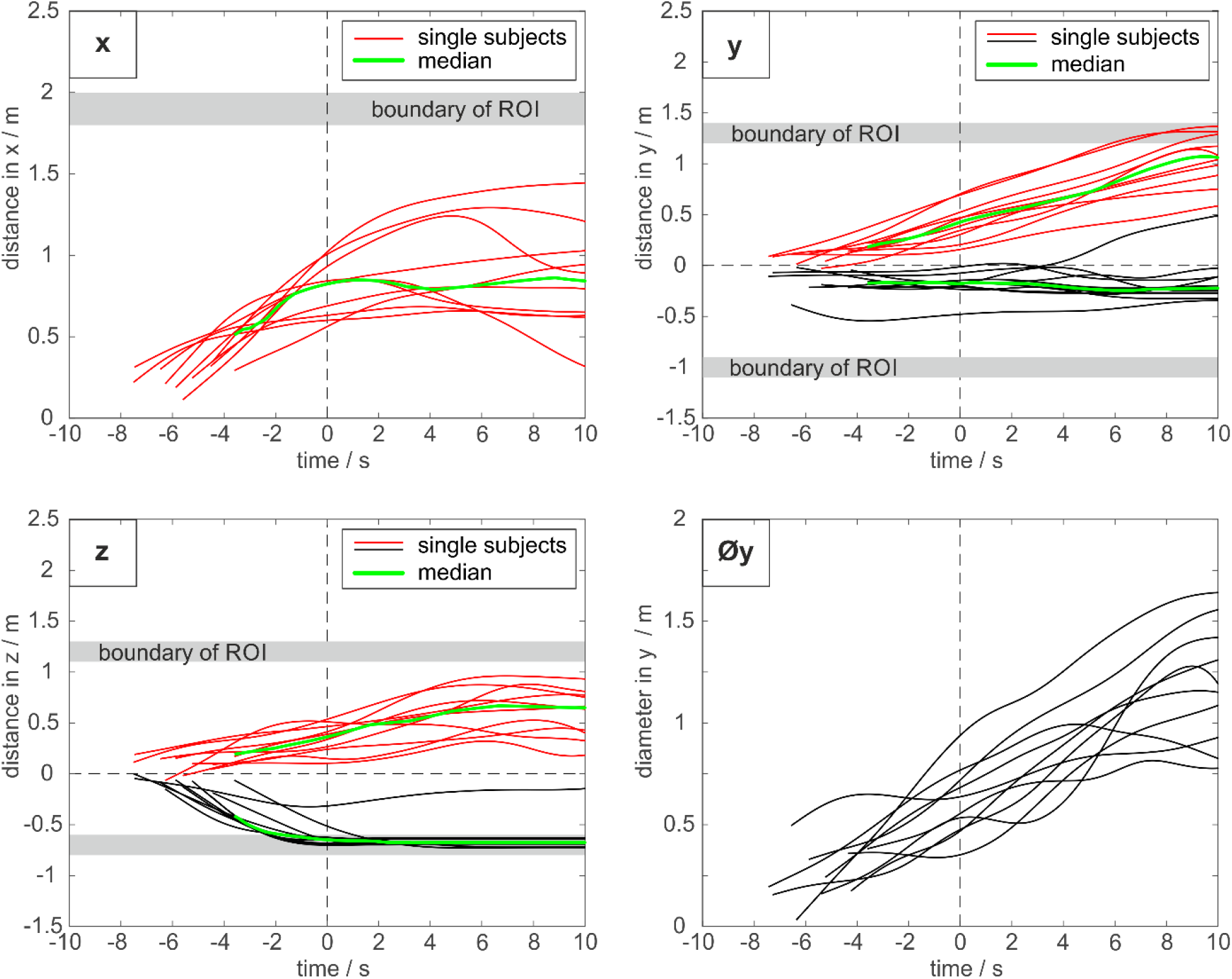
Traces of all subjects for all dimensions (a: x-dimension (front), b: y-dimension (left-right), c: z-dimension (Up-down) and d: diameter of the y-dimension) for the T+ task. The 0 point in the time-scale refers to the end of the task. The green lines show the median. The corresponding graphs for the other tasks are provided in the supplemental material.

